# Patient and Public Involvement in designing a clinical trial for the surgical management of persistent Post-Infectious Olfactory Dysfunction

**DOI:** 10.64898/2026.09.08.26362544

**Authors:** Aagat Sharma Khatiwada, Laurie R Springford, Christine Kelly, Alfonso Luca Pendolino, Lynne Corner, Peter J Andrews

**Affiliations:** Ear Institute, University College London (UCL), London, UK; St George’s University Hospitals NHS Foundation, London, UK; CKOS, Andover, UK; Department of ENT, Imperial College Healthcare NHS Trust, London, UK; VOICE, Newcastle University, Newcastle, UK; Department of ENT, Royal National ENT & Eastman Dental Hospitals, London, UK

**Author notes:** Correspondence to: Mr Aagat Sharma Khatiwada, MRCS(ENT) MBChB(Hons), Ear Institute, University College London (UCL), London, UK WC1X 8EE. **Conflict of Interest:** The authors declare that they have no conflict of interest.

**Keywords:** patient and public involvement, post-infectious olfactory dysfunction, functional septorhinoplasty, randomised controlled trial design, GRIPP2, health inequalities

## Abstract

**Introduction:** Persistent post-infectious olfactory dysfunction (PIOD) has limited effective treatments. In developing a randomized controlled trial (RCT) to evaluate a novel surgical intervention for persistent PIOD, embedding Patient and Public Involvement (PPI), a core NIHR requirement in research, was essential.

**Objectives:** To outline the PPI programme for our proposed RCT, in line with UK Standards for Public Involvement. The objective of the PPI programme was to design a patient-centred, inclusive, and ethically sound trial.

**Methods:** The PPI programme included an online survey, online discussion forums, and focus groups.

**Results:** 63 survey responses, 2 online discussion forums, and 2 focus groups provided PPI input on control arm acceptability, participant recruitment and retention strategies, participation burden, and trial outcome selection, directly shaping the trial protocol.

**Conclusions:** This study demonstrates how robust PPI in trial design ensures that clinical trials remain responsive to the true priorities of the patient by aligning scientific rigor with lived experience.

## INTRODUCTION

Olfactory dysfunction (OD) represents a significant burden for the general population, with over 20% affected.^1^ Post-infectious olfactory dysfunction (PIOD) accounts for 18-45% of OD.^2^ The COVID-19 pandemic saw a marked increase in PIOD, occurring in up to 48% of patients in the earlier variants, and brought olfaction in sharp focus within medical research.^3^ Long-term follow-up data indicate that approximately 8% of individuals with COVID-19-related olfactory dysfunction (C19OD), a subset of PIOD, experience persistent symptoms at 3 years. This represents up to 400,000 new annual cases in the UK during the pandemic period.^4^ Yet, as public interest in the COVID pandemic and its consequences wanes, this large group of patients suffering from persistent PIOD is at risk of being forgotten.

OD affects multiple domains of someone’s life, including nutrition, hedonic pleasure, personal safety (e.g. detection of smoke, gas, or spoiled food), and social functioning. It is associated with a significant mental health burden, including depression, and a diminished quality of life.^5,6^ One smell-loss sufferer described it as “living in a cardboard box…disconnected from the pleasures of the world,” and another said “it is so debilitating and causes long-term anxiety and feelings of loss.”

Despite this growing prevalence and impact, effective treatments for OD remain limited and experimental. Olfactory training (OT) is currently the only therapy supported by Level 1 evidence. Yet, OT is effective in only around a quarter of patients with persistent PIOD, partly owing to poor compliance.^7,8^ A majority of patients are therefore left with ongoing functional impairment, contributing to the ubiquitous patient perception of OD being neglected or disregarded by clinicians.^9^ Indeed, many patients with persistent PIOD live with an impairment that is inadequately addressed in clinical practice and within research priorities at national level, despite the substantial burden of disease.

Functional septorhinoplasty (fSRP) involving the widening of the internal nasal valve using spreader grafts has been shown to increase airflow to the olfactory cleft, and represents a novel, mechanistically informed, intervention for persistent PIOD. A 2023 proof-of-concept (PoC) study was the first to demonstrate that fSRP can lead to clinically meaningful improvements in olfactory function in patients with persistent PIOD.^10^ A subsequent PoC study (2025) focusing on persistent C19OD confirmed these findings, demonstrating improvements exceeding the minimal clinically important difference (MCID) compared with patients continuing OT.^11^

To delineate the exact mechanistic basis for these results, we are planning a randomised controlled trial to evaluate the clinical effectiveness of fSRP for persistent PIOD. The study is planned to incorporate a high-fidelity surgical placebo control arm (septoplasty without spreader grafts), removing the key mechanism (spreader graft insertion) responsible for further increasing olfactory airflow above normal levels. Olfactory mucosa biopsies and functional MRI scanning of patients are also planned to investigate the mechanistic pathways involved in persistent PIOD and olfactory recovery after fSRP.

Patient and public involvement (PPI) in clinical trials is a core expectation of National Institute of Health and Care Research (NIHR), and a requirement for research funding. The NIHR defines PPI in research as “research being carried out ‘with’ or ‘by’ members of the public rather than ‘to’, ‘about’ or ‘for’ them.”^12^ High quality PPI improves the relevance and quality of research by involving members of the public in the planning, acceptability, study design, recruitment, outcomes and dissemination.^13^ Consequently, the inclusion of PPI in research, as well as its reporting has increased over time, now embedded as key feature of research culture.^14,15^ Here we report the PPI initiatives undertaken as part of the design of our clinical trial investigating the effects of fSRP on olfactory function in patients with persistent PIOD.

## OBJECTIVES

The main objectives of the PPI for this study are as follows: to ensure the trial design is grounded in the priorities and lived experience of people with PIOD; to improve the quality, acceptability, accessibility, and ethical conduct of the trial; and to maximise inclusive reach into underserved communities affected by smell loss.

Our PPI programme has been developed against the UK Standards for Public Involvement (Inclusive Opportunities, Working Together, Support and Learning, Governance, Communications, and Impact).^16^ The report has been presented using the internationally agreed standardising for report of PPI in healthcare research, GRIPP2.^17^ The report pertains to the PPI activities so far undertaken, and a summary of the planned PPI involvement throughout the lifecycle of the study is presented.

## STUDY DESIGN

We used a sequential mixed-methods PPI design, combining an online survey, two online discussion forums, and two focus group meetings, to inform our study design. Each PPI activity is described in detail below.

## METHODS

### Survey

A plain-English summary of our proposed study was drafted, adapted to Google Forms, and distributed alongside a survey of 10 questions. Google Forms was felt to be the most accessible platform, and the survey itself was amended following feedback from CK, who runs the popular online smell loss platform, Chrissi Kelly on Smell (CKOS), formerly AbScent.

The survey was divided into the following sections: 1. Understanding the study 2. Importance and relevance 3. Acceptability of surgery for smell loss 4. Views on the study design 5. Random allocation and chance of receiving the intervention 6. Burden of tests and follow-up 7. Biopsy 8. What matters to you (study outcomes) 9. Overall willingness to take part, and 10. Optional follow-up involvement.

The survey used a mixed-method design combining quantitative rating scales (Likert-style rating scale, multiple choice checkboxes etc.) with qualitative open-ended feedback questions. We estimated between 3-5 minutes for completion of the survey.

The survey was advertised on CK’s online platforms including the CKOS Network of 5000 people suffering from olfactory disorders, Covid Facebook group of 32,300 members, Facebook parosmia group of 21,100, the generic Facebook Abscent group of 7,400 members, and CK’s online newsletter distributed to 5,500 members, with an open rate of 44%.

### Discussion forum

Following the survey, two online discussion forums were organised through CK’s platforms. CKOS/Abscent runs a regular monthly online meeting normally attended by an unselected group of smell loss sufferers. Our study was highlighted as a key agenda item for their February and April 2026 meetings, and a member of the research team attended the meeting to discuss the study.

Two online discussion forums centred around the trial were held. They were conducted on Zoom, and moderated by CK.

### Focus group meetings

Participants from the online discussion forums who expressed an interest in joining the core PPI group were invited, via email. Two focus group meetings were arranged with a member of the research team, CK, and PPI participants. Focus group meetings were held on Teams. Study related materials, including plain-English summary, were distributed beforehand.

To guide focus group discussions, qualitative interview-style questions and prompts were aligned with the NIHR’s UK Standards for Public Involvement in Research. Following feedback from the wider research team, including CK, the final agenda was structured into three sections: (1) Opening summary and study design overview, (2) Acceptability of the control group, and (3) General questions on study design based on NIHR PPI standards.

Participants consented to anonymised quotation for this publication.

## RESULTS

### Survey

A total of 63 responses was received. 100% of survey respondents agreed that there was an urgent need for research into smell loss. 92% of respondents viewed surgery as an acceptable treatment option for olfactory loss (with 3% deeming it unacceptable), and 84% expressed willingness to participate if eligible. In addition, 92% endorsed the inclusion of an olfactory mucosa biopsy. Participants identified the ability to smell everyday odours as the primary outcome measure of importance, followed by quality of life, emotional well-being, and taste restoration.

Feasibility assessments regarding trial design revealed nuanced participant perspectives. While 46% were comfortable using septoplasty as a control procedure, 27% felt its inclusion reduced the trial’s acceptability (the rest were undecided). Concerns focused on undertaking operative risks without a proportionate likelihood of benefit. Specific concerns centred on scarring after control surgery, and its implications on the feasibility of future fSRP. As a way of mitigating concerns around the control arm, participants strongly advocated for a post-trial crossover option. Regarding random allocation, preferences were divided equally between 2:1 (higher chance of receiving the intervention) and 1:1 randomization (38%, n=24/63 each, rest unsure). Proponents of 2:1 highlighted that the higher probability of receiving the intervention better respected the risks and burdens of undergoing surgery, whereas those favouring 1:1 prioritized scientific and methodological rigour.

The survey also stimulated discussions on online platforms. Qualitative feedback included concerns regarding invasiveness (“it sounds invasive”) and control arm placement (“I would hate being in the control group”). Yet participants frequently expressed feeling “desperate” and “dismissed,” with one participant saying “I would do anything to get my smell back.” Equally, public engagement highlighted strong endorsement of novel research (“understanding of anosmia cannot move forward unless new initiatives are explored”) and mechanistic curiosity (“I need to know how a slight increase in airflow can restore sense of smell”). Alongside this, concerns about equitable access were raised: “everything appears to be a postcode lottery to me,” or someone expressing having to “fight a battle to get past the triage system.” To directly address these questions and refine the protocol, subsequent online discussion forums were organised.

### Discussion forum

Each online discussion forum was attended by up to 12 participants and lasted approximately one hour. All participants experienced olfactory loss, and represented a diverse range of underlying aetiologies (e.g., post-infectious, COVID-19-related, traumatic, and chronic rhinosinusitis). While the majority of participants were UK-based, international representation included individuals from the United States.

The discussion forum started with an overview of the proposed trial that directly addressed the concerns that had previously been raised on the survey and online platforms. Discussion centred on the logistical and mechanistic rationale for the control arm and control procedure, as well as hypotheses surrounding how fSRP might improve smell loss.

Concluding verbal feedback demonstrated that participants had gained a clearer understanding of the study design, appreciated the candour around clinical evidence gaps, and reiterated the necessity of research in this area. These discussion forums facilitated direct public engagement and effectively addressed baseline concerns. Crucially, engagement at the forums served to recruit a dedicated core PPI group for the remainder of the study.

### Focus group meetings

Following the online discussion forums, four participants contacted the research team and CK, and the core PPI advisory group was formed with these participants. Two 1-hour focus group meetings were conducted online via Microsoft Teams, scheduled one week apart. All PPI representatives were UK-based and had lived experience of post-viral or COVID-19-related olfactory dysfunction. The meetings were attended by a research team member, CK, and 2-3 PPI representatives. The focus group discussions were thematic, evaluating key trial dimensions, as follows:

### Acceptability of septoplasty/control procedure

Detailed explanations of the procedural steps and mechanistic rationale behind the intervention (fSRP) and control arms (septoplasty) were provided. The research team detailed the hypothesis of improved nasal airflow and olfactory function, and reiterated the ambiguity around the therapeutic impact of either surgery on post-viral olfactory loss.

Overall, participants demonstrated high acceptability toward the surgical control arm, recognizing its scientific necessity. However, acceptability was strongly contingent upon incorporating a post-trial crossover provision, enabling control participants to receive SRP if therapeutic efficacy is established. This was consistent with the survey findings. When presented with a hypothetical control arm of olfactory training (OT) alone, participants rejected this in favour of a surgical control arm, noting that most eligible individuals would have already trialled OT and would prefer an active surgical intervention despite uncertain efficacy. To fully convey the risks associated with both the intervention and control procedures, the group recommended using accessible visual media (diagrams, videos, and animations), and proposed a peer-support’buddy scheme,’ pairing trial candidates with individuals who have previous lived experience of these procedures.

### General Questions on Study Design based on NIHR PPI Standards

Reach and inclusion: The group explored strategies to optimize trial reach and equitable access, particularly among underserved populations. To engage younger audiences, targeted outreach across diverse social media platforms was recommended. On the other hand, to improve the inclusion of older demographics, marginalised groups (e.g. ethnic minorities), or non-digital users, traditional pathways such as direct GP email communications and physical advertisements on hospital notice boards were advocated. Furthermore, participants emphasized leveraging the communication networks of participating research centres to maximize community awareness.

### Burden of participation

The proposed follow-up schedule at 3, 6, and 12-months post-operation was fully acceptable to the group. Any travel burden was advised to be mitigated with expense reimbursement.

Olfactory mucosa biopsies were largely acceptable, with most participants expressing no hesitations. However, a specific safety concern was considered: whether harvesting tissue from individuals with preexisting, chronic smell loss might further compromise their remaining olfactory function. The research team presented safety evidence from existing literature and pilot study data demonstrating no post-biopsy deterioration, which satisfied the group, provided that prospective trial participants receive a detailed, explicit disclosure of biopsy risks during the informed consent process.

Assessment of the functional MRI scanning protocol yielded no objections, though the group emphasized the necessity of a formal protocol to manage incidental imaging findings.

Regarding participant retention, to mitigate dropout risks, the group recommended assigning a dedicated research nurse or liaison to maintain proactive communication with participants between surgery and scheduled follow-up visits, ensuring ongoing engagement and support.

Outcomes that matter to patients: Participants supported the use of objective olfactory testing with Sniffin’ Sticks as the primary outcome measure. To mitigate the potential discrepancies between psychophysical test scores and patients’ subjective perceptions of smell, they recommended incorporating visual analogue scales and validated quality-of-life questionnaires, along with questionnaires to capture the specific impacts of parosmia and phantosmia. Additionally, the group emphasized the profound impact of olfactory loss on flavour perception and enjoyment of food, and recommended questions to assess flavour perception and food enjoyment.

### Follow-up length and support

The group concurred that a 12-month follow-up period represents a pragmatic design choice aligned with current literature. However, they recommended obtaining participant consent at trial entry for optional, extended follow-up. This approach enables future cohort re-evaluation beyond 12 months without delaying primary outcome reporting.

### Governance and ongoing involvement

All focus group representatives agreed to join the permanent core PPI advisory group, committing to ongoing collaboration and patient oversight throughout the remaining lifecycle of the trial.

## DISCUSSION

Our comprehensive Patient and Public Involvement (PPI) initiative comprised an online survey, online discussion forums, and focus groups. These PPI activities provided critical lived-experience insights aligned with the NIHR Standards for Public Involvement, and are reported here in accordance with the GRIPP2 checklist.^17^ They led to significant impact on the trial design, a summary of which is provided in Table 1.

**Table 1:** Summary of PPI feedback, source of feedback and trial protocol change directly resulting from the feedback.

| PPI feedback | Source of feedback | Resulting protocol change |
| --- | --- | --- |
| *Choice of comparator arm: OT alone rejected, septoplasty endorsed* | Survey, focus groups | Septoplasty retained as comparator in control arm |
| *Offer of intervention to control participants at study-end* | Survey, discussion forums, focus groups | fSRP to be offered to control arm participants at study end if benefit demonstrated |
| *Randomisation protocol: equal support for 1:1 and 2:1 (intervention) randomisation* | Survey | 1:1 randomisation retained |
| *Buddy-scheme: Optional peer support scheme pairing participants with past septoplasty/fSRP patients* | Focus groups | Peer support scheme built into study protocol |
| *Outcome measures: subjective outcome measures recommended, including inclusion of qualitative olfactory dysfunction* | Focus groups | Inclusion of visual analogue scale of smell function, and assessment of parosmia and phantosmia within outcome measures |
| *Olfactory mucosa biopsy safety:* | Focus groups | Inclusion of olfactory mucosa |
| detailed discussion of olfactory mucosa biopsy safety prior to consenting |  | biopsy safety to patient<br>information leaflets, discussion of safety data built into consent pathway |
| *Recruitment and dissemination strategy and steps to optimise inclusivity* | Focus groups | Expansion of recruitment and dissemination strategy to include GP mailshots, hospital board ads, radio ads, local hospital and community network ads, and diversification of social media platforms |
| *Retention strategy:* research nurse/liaison for continued participant engagement | Focus groups | Research nurse liaison with scheduled interim retention calls between study visits integrated into protocol |

### PIOD: a persistent and under-recognised problem

First-hand participant testimonials underscored a profound sense of despair and frustration at low levels of awareness of OD within the healthcare community, a paucity of effective treatment options, and a perceived lack of dedicated research and clinical prioritisation, echoing previously reported barriers to care.^9^ Plainly, for many patients who have contributed to this work, PIOD is a persistent, disabling, and in their own words ‘forgotten’ condition that requires high quality research driven treatment options. Since the media focus from COVID anosmia has moved on, this subset of the population risk being left with limited treatment options and no evidence base to turn to. From the patient testimonials, the desperation that smell loss sufferers feel became apparent. The proposed surgical trial, therefore, was met with strong endorsement: 100% of survey respondents concurred with the need for urgent research into smell loss, and 92% deemed surgical intervention acceptable despite inherent procedural risks.

To address the specific concerns raised during our PPI engagement such as general anaesthetic risks, procedural complications, and scarring, the research team plan to develop accessible visual information sheets, multimedia material (including videos comparing fSRP and septoplasty), and explicit disclosures regarding existing evidence gaps. Specific material detailing the safety profile of olfactory mucosa biopsies will directly address the concerns raised regarding the risk of further olfactory compromise after biopsy. Additionally, a ‘buddy scheme’ will allow prospective participants to gain an unbiased perspective from individuals who have previously undergone septoplasty or fSRP.

To optimize acceptability of the trial design, several key structural refinements were instituted. The study now guarantees post-trial crossover access to SRP for control-arm participants should the intervention demonstrate efficacy. Regarding allocation, PPI insights confirmed the acceptability of a 1:1 randomization scheme. While 1:1 and 2:1 randomisation received equal support in the survey, the 1:1 randomisation was opted for the preservation of methodological rigour, a decision which was subsequently endorsed in the focus groups. Furthermore, participants viewed a surgical control (septoplasty) more favourably than a non-surgical control arm with olfactory training alone.

To mitigate participant attrition, a dedicated research liaison will maintain regular proactive contact with participants between trial visits. Furthermore, a standardised pathway will ensure that incidental findings on MRI scanning are acted upon.

### Reach, equity and inclusion

Anxiety over equitable access to the trial was frequently shared during the PPI process, and is evident from the patient testimonials described above. To overcome geographic and socioeconomic disparities in access to specialized NHS smell services, trial recruitment has been distributed across four sites in London, the Midlands (Birmingham), and the North (Manchester). Such a distribution will ensure equitable access to diverse communities throughout the UK. Inclusivity will be enhanced further by multi-channel advertising, utilizing social media for younger demographics, and direct GP outreach networks, hospital board displays, and radio ads for non-digital, marginalised groups, and older populations. Patients will be supported by built-in travel expense reimbursement as a means of reducing barriers to participation in the study.

Additionally, we have leveraged the Voice Global (VOICE) platform to enhance reach and inclusivity, and reflect the multisite nature of the trial.^18^ VOICE is an established, patient and public community of approximately 4,000 members across the UK. Its digital infrastructure on a national scale provides a mechanism to coordinate public involvement, sharing research documents and knowledge exchange on the topic, supporting genuine co-production and the meaningful involvement of the public on this issue. Importantly, the VOICE platform will be leveraged to optimise participant recruitment and trial outcome dissemination, enhancing targeted outreach to under-represented groups throughout the lifecycle of the trial.

### What matters to patients?

While PPI participants endorsed Sniffin’ Sticks as the primary objective endpoint, the outcome measures were expanded to capture subjective lived experience.^19^ Secondary measures therefore incorporate Visual Analogue Scales for subjective smell perception, validated olfactory quality-of-life instruments, specific metrics evaluating dysosmia (parosmia and phantosmia),^20,21^ and dedicated instruments measuring retronasal flavour perception and food enjoyment.^22^

Finally, while the pragmatic 12-month follow-up schedule was retained, initial trial consent will now include an optional provision for extended long-term cohort tracking beyond the primary outcome deadline.

### PPI plan for the remainder of the trial

All 4 focus group representatives have transitioned into a permanent core PPI advisory panel, which will convene regularly for focus group meetings once every 2 months. Beyond the initial setup phase, public involvement remains embedded throughout the trial lifecycle. Guided by the VOICE Global platform, the Core PPI Group will actively support participant recruitment and messaging, review the peer support ‘buddy’ scheme, and gather interim feedback on consent, randomisation, and perioperative pathways. As the trial progresses, contributors will evaluate follow-up acceptability, refine retention strategies, and monitor adverse experience feedback reported via patient forums. In the final phase, public contributors will collaborate with clinical and statistical teams to interpret findings, co-produce lay summaries, and drive dissemination across public channels alongside academic publication. Throughout, all PPI input will be evaluated against the UK Standards for Public Involvement, ensuring transparent feedback loops and continuous impact reporting. In terms of PPI governance, a named PPI lead will report directly to the Trial Management Group, alongside formal representation on the Trial Steering Committee. Public contributors have so far been compensated for their involvement in PPI activities, and will continue to be paid for their time in line with current NIHR guidance.

### Reflections and Limitations

Reflecting on the PPI process highlights both procedural challenges and valuable insights gained throughout the engagement. Initially, variation in participants’ understanding of the PPI framework created confusion, with some mistaking the invitation for PPI involvement as recruitment into the clinical trial itself. This early communication gap was successfully addressed through iterative refinements guided by our patient advocacy lead, CK.

A further limitation relates to the use of online medium for all PPI activities. Relying exclusively on digital platforms risks excluding populations with limited digital literacy or access, as well as those who do not engage with specialised online support networks such as AbScent. This selection bias was reflected in our focus group, which included participants of a specific demographic, to the exclusion of others. We therefore consider this one of the significant gaps in the current PPI program with reference to the UK Standards for Public Involvement.^16^ Steps to mitigate this selection bias include recruiting patient representatives directly through hospital outpatient clinics, community healthcare settings and through the VOICE partnership engagement.

Notwithstanding these limitations, the dedication and generosity of the participants proved invaluable. Representatives voluntarily contributed significant time and energy to refine the trial protocol, demonstrating a deep commitment to advancing research in olfactory dysfunction. Their shared enthusiasm and lived-experience perspectives have ultimately reinforced the research team’s resolve to address critical evidence gaps in post-viral smell loss.

## Data Availability

All data produced in the present study are available upon reasonable request to the authors

